# Death, Culture, and Conflict: A Qualitative Study on Sociocultural Practices and Their Implications for Maternal and Perinatal Death Surveillance in Eastern Democratic Republic of Congo

**DOI:** 10.64898/2026.07.22.26358727

**Authors:** Meighan Mary, Christine Chimanuka Murhima’alika, Christian Chiribagula Zalinga, Christian Mugisho Byamungu, Pacifique Mwene-Batu, Emilie Grant, Rosine Bigirinama Nshobole, Gaylord Ngaboyeka, Salomine Ekambi, Hannah Tappis, Ghislain Bisimwa Balaluka

## Abstract

**Background:** Death is a social and cultural phenomenon whose meaning shapes how and whether losses are mourned, disclosed, and reported. These dynamics have direct implications for maternal and perinatal death surveillance and response (MPDSR), yet remain understudied, particularly in humanitarian contexts.

**Methods:** This phenomenological qualitative study was conducted in two conflict-affected health zones in Eastern Democratic Republic of Congo. In-depth interviews (n=50) were conducted with bereaved family members of maternal or perinatal deaths, community leaders, and health providers to understand the socio-cultural practices surrounding death and the factors influencing MPDSR. Interviews were transcribed in French and analyzed using inductive thematic content analysis.

**Results:** Four themes characterized the socio-cultural practices surrounding maternal and perinatal deaths: burial practices, mourning and bereavement traditions, rationale for these practices, and the impact of insecurity on customs. Burial and mourning practices differed markedly by type of death, with stillbirths and neonatal deaths accorded significantly less social recognition than maternal deaths. Deaths were commonly attributed to witchcraft or spiritual causality, or blame directed at mothers, husbands, and health providers. Active conflict further disrupted customary practices and eroded community trust. Collectively, these dynamics inhibit disclosure and reporting of deaths, undermining MPDSR case identification.

**Conclusion:** Effective MPDSR in conflict-affected settings requires culturally responsive adaptation, community involvement in case identification, and trust in health sector actors. By documenting specific actors involved in burials, variations in burial and mourning practices, and how conflict changes socio-cultural practices, findings offer actionable entry points for strengthening MPDSR in conflict-affect health zones in Eastern DRC.

## Introduction

Maternal and perinatal deaths remain pressing public health challenges, with the highest burden in low- and middle-income countries (1). Beyond their medical causes, these deaths are embedded in social and cultural norms that shape how they are understood, experienced, and reported. In many settings, community beliefs about stillbirth, neonatal deaths, and maternal deaths influence whether families disclose these losses, how they mourn, and whether they engage with formal health structures (2–4).

Cultural and religious beliefs strongly shape how stillbirths, neonatal deaths, and maternal deaths are understood. Stillbirth, for example, is often viewed as an act of divine will, with the stillborn child regarded as a stranger both socially and spiritually, despite the trauma to parents (3,5) or as part of the natural cycle of life (5–7). Neonatal mortality, by contrast, is typically recognized as the passing of a baby that was born alive and is often interpreted as medical failures, particularly when sudden or unexpected. In some contexts, such losses may also be attributed to mystical causes such as witchcraft (8,9). Similar to stillbirths, neonatal deaths can be explained within religious or spiritual lenses, with many communities ultimately framing them as acts of divine will (2,10,11). Maternal mortality is similarly shaped by cultural and spiritual interpretations, particularly under socially stigmatized circumstances such as pregnancy outside marriage or unsafe abortion, where deaths may be understood as spiritual punishment or the result of occult forces (3,7).

These socio-cultural interpretations not only affect the grieving process but also have direct consequences for health information systems, particularly Maternal and Perinatal Death Surveillance and Response (MPDSR)(12). Families may choose silence to avoid stigma, turning instead to traditional chiefs, pastors, or healers rather than formal health facilities (3,7,13,14). While deeply rooted in local belief systems, these practices conflict with official reporting procedures and contribute to underreporting. Deaths concealed due to stigma, or reported only within spiritual or traditional domains, bypass the MPDSR cycle of notification, review, and response, leading to gaps in critical data and weakening the system’s ability to identify preventable factors and recommend interventions (15,16). In this way, cultural understandings of death shape grieving practices and can limit accountability mechanisms intended to reduce maternal and perinatal mortality.

Cultural practices surrounding maternal and perinatal deaths add another layer of complexity. For example, some mothers of stillbirths may forego mourning or bereavement periods (7,17), while those in other cultures may consider stillbirth funeral rituals as essential to uphold cultural ties and to accompany the deceased to their final resting place (17). Neonatal and maternal bereavement practices can also vary widely, from multi-day mourning ceremonies to restrictions on mothers’ participation, or rituals intended to prevent spiritual harm (11,17,18). While these practices are important for families, they may contribute to delayed notification or underreporting. In conflict-affected settings, where displacement a disrupt established norms, barriers to reporting are worsened by mistrust of health structures and heightened stigma (16). These challenges conceal the actual burden of preventable mortality and limit the capacity of surveillance systems to respond effectively.

In the Democratic Republic of Congo (DRC), several United Nations agencies and partners collaborated to support the Ministry of Health in developing MPDSR guidelines and reporting tools (19). The DRC national guidelines outline expectations for MPDSR implementation, including both facility and community level approaches. However, as documented in other humanitarian settings (20–22), awareness of this guidance and implementation is weak or absent in Eastern DRC (23). This study is part of a portfolio of work led by the EQUAL Research Consortium (24) to strengthen MPDSR in conflict-affected zones in North and South Kivu Provinces of DRC. The study aims to understand the socio-cultural practices surrounding death and the factors influencing the reporting of maternal and perinatal deaths in Mweso (North Kivu) and Mulungu (South Kivu) health zones, where humanitarian crises and cultural diversity make MPDSR implementation particularly complex and challenging. Examining how deaths are interpreted within communities provides critical insights into the barriers and opportunities for strengthening death reporting and improving maternal and newborn health outcomes.

## Methods

### Study Design

A phenomenological qualitative study was conducted to understand the socio-cultural practices surrounding death and how communities conceptualize and make sense of maternal and perinatal deaths in crisis-affected contexts. In-depth interviews (IDIs) were undertaken with 1.) bereaved family members of maternal and perinatal deaths, 2.) community leaders, and 3.) health providers. The Johns Hopkins Bloomberg School of Public Health Institutional Review Board (21935/MOD3503) and the Université Catholique de Bukavu Institutional Health Ethics Committee (UCB/CIES/NC/022/2022) approved the research. Data collection was undertaken in 05/12/2022 – 23/12/2022 in South Kivu Province and 28/06/2023 – 15/07/2023 in North Kivu Province in compliance with the Declaration of Helsinki (25) and all local regulations and institutional policies.

### Study Location

This study was conducted in two health zones in Eastern Democratic Republic of Congo: Mweso Health Zone in North Kivu Province, Masisi Territory, and Mulungu Health Zone in South Kivu Province, Mwenga Territory. Both regions have been affected by prolonged armed conflict (26) and were purposively selected to capture the challenges of implementing the MPDSR in varying humanitarian contexts. For more than three decades, the Eastern DRC has been plagued by persistent insecurity, characterized by repeated population displacements, a weakened health system, and limited access to healthcare services (26). In this context, access to healthcare for women and children remains particularly difficult, and a significant proportion of births, as well as maternal and perinatal deaths, remain unreported (27).

The Mweso Health Zone (administrative district) comprises 23 health areas (sub-district) and a general referral hospital. It is characterized by multiple low-to medium-intensity conflicts due to the strong presence of armed groups, leading to mass population displacement (26). It is also supported by humanitarian organizations that facilitate free access to healthcare in health facilities. In contrast, the Mulungu rural health zone is geographically isolated off due to a lack of passable roads and insecurity. It comprises 20 health centers and a general referral hospital. It has limited access to health services due to inadequate infrastructure, as observed in many fragile and conflict-affected rural settings (26).

**Figure 1:**
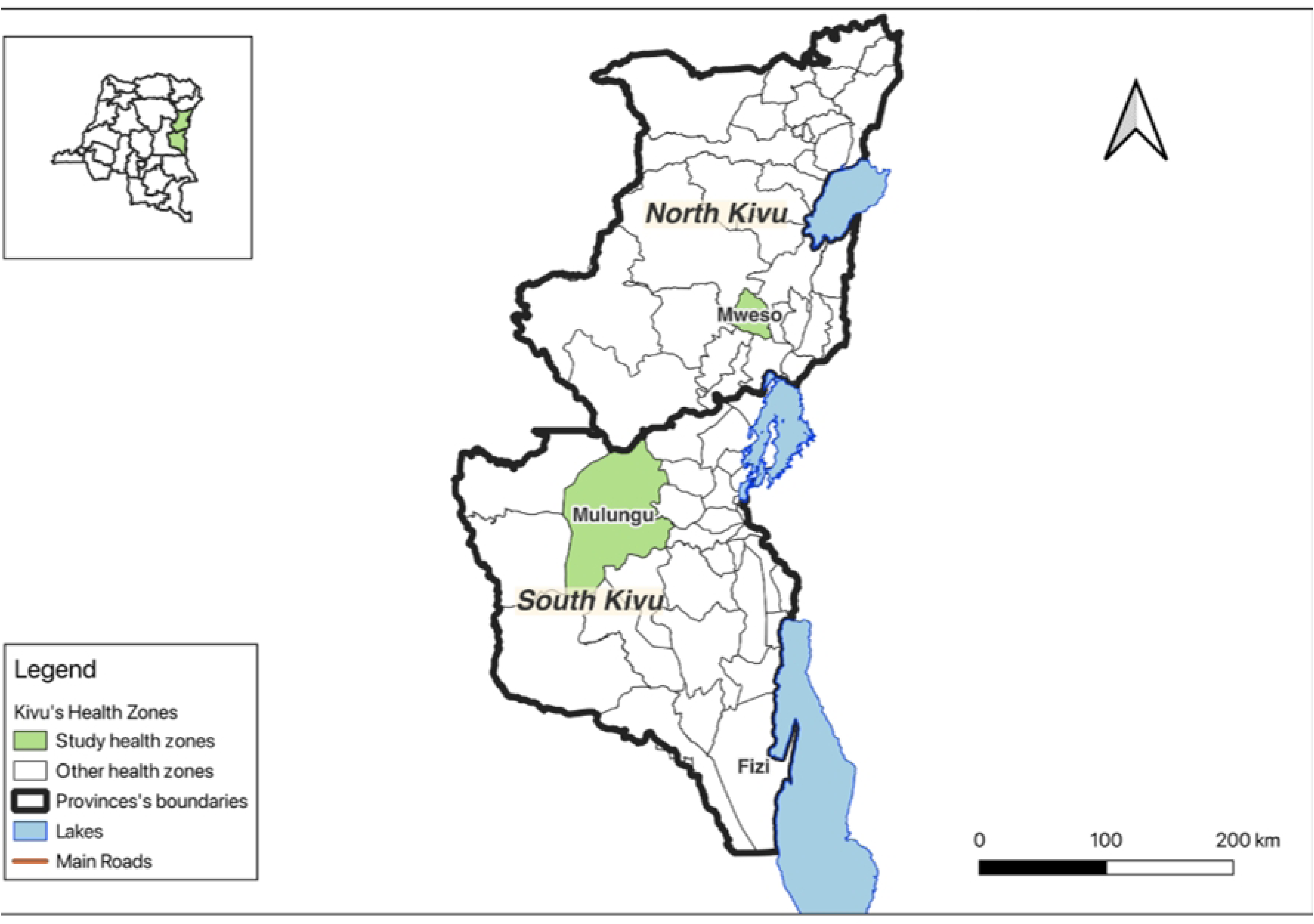
Map of Study Sites

### Study Sample

A purposive sample of bereaved family members, community leaders, and health providers were selected for the IDIs. In consultation with community health committees and the community health worker (CHW) networks in Mweso and Mulungu, villages were identified where families who recently experienced a stillbirth, maternal or neonatal death (in the prior three months). Within these villages, a bereaved family member (e.g., mother or husband) and a community leader were invited to participate.

Selection of participants was designed to ensure a diversity of perspectives. Bereaved family members of maternal and perinatal deaths were selected according to the type of death (stillbirth, maternal or neonatal death), their status within the conflict-affected zone (host community or refugee/displaced person), and their proximity to the death and/or involvement in the birth. Bereaved family members and leaders were excluded if they did not reside in Mweso or Mulungu health zones.

Within each health zone, health providers were selected from the sole tertiary hospital and two randomly selected health centers that reported a maternal or perinatal death in the 12 months prior to data collection. Health providers were selected to capture perspectives from diverse cadres including obstetricians, nurse/midwives, and CHW. Only participants 18 years or older were eligible to participate. Health providers were ineligible if they were not involved in a maternal or perinatal death case in the prior 12 months or at their current location of employment (health facility).

With this participant selection approach, a sample of 12 bereaved family members, 4 community leaders, and 9 health providers was estimated to achieve saturation of themes in each health zone, for a total of 50 participants.

### Data Collection

In advance of data collection, a team of three qualitative interviewers with expertise in public health and social science underwent a five-day training led by the Université Catholique de Bukavu and Johns Hopkins Bloomberg School of Public Health research teams. The training focused on the study design and methodology, research ethics, and sensitive qualitative interviewing techniques, and included time for thorough piloting of the interview guides.

Once a community where a family experienced a maternal or perinatal death was identified, a CHW accompanied the study team to visit the family and invite a member close to the deceased to participate in an IDI. If interested in participating, a study team member confirmed their eligibility and obtained written informed consent in advance of data collection. Interviews were conducted in French or Swahili based on preference of the interviewee using a piloted semi-structured interview guide, in a location of the participant’s choosing that could ensure visual and auditory privacy for the duration of the discussion. IDIs lasted 30-60 minutes. IDIs with community leaders were conducted in the same villages using the same data collection procedures.

At each selected health facility, the interview team thoroughly introduced the study to all health personnel in the maternity and neonatal departments. Recruitment was managed with sensitivity and transparency to avoid the suspicion of blame when seeking to interview a health provider that participated in a maternal or perinatal death case. When a health provider indicated interest in participating, the study team confirmed eligibility, obtained written consent, and conducted an IDI in a private office or confidential location of their choosing.

### Data Analysis

All IDIs were translated in French, transcribed verbatim, and uploaded to Dedoose (version 8.3) qualitative software. Thematic content analysis was conducted by MM and CCM and employed an iterative inductive coding process to develop the qualitative codebook based on emergent themes related to sociocultural beliefs and practices surrounding death. Thematic tables were constructed to elucidate triangulated perspectives from bereaved family members, leaders, and health providers findings by type of death (maternal death, neonatal death, or stillbirth).

### Reflexivity

Reflexivity was practiced during data collection and analysis (28). All Interviewers (two women and one man) underwent training in interpersonal and cultural reflexivity, with emphasis on recognizing and addressing gender, social, and power dynamics. They participated in regular debriefing to reflect on how their positionality, personal assumptions, and interactions with participants might have influenced data collection. Interviewers had no prior relationship with the participants.

The practice of reflexivity was continued by co-author involvement. Co-authors represented the DRC and the United States and co-PI, and analysts MM and CCM are ci-gendered women with personal birthing experience. Weekly meetings were conducted to review coding decisions, discuss emerging themes, and reflect on how team members’ perspectives might have shaped interpretation. Discrepancies were addressed through reflexive discussion and resolved with consensus to strengthen analytical rigor.

## Results

A total of 49 participants shared their perspectives on socio-cultural practices surrounding maternal and perinatal mortality. Table 1 describes key participant characteristics. Among bereaved family members of maternal and perinatal deaths, 70% were mothers of stillborn or neonatal deaths and 30% were fathers or husbands of women who passed in pregnancy or postpartum. Most bereaved family members (78%) were 20-40 years old and approximately one third (35%) were internally displaced persons (IDP) or refugees. Education level varied, yet almost one fifth had received no education (17%). Community leaders were primarily male (75%) and represented the IDP/refugee and host communities equally. In alignment with the sampling strategy, health providers were primarily nurses or midwives (67%), followed by CHWs (22%) and obstetricians (11%). Over half of health providers were female (55%) and had worked 1-5 years at the selected health facility (61%).

**Table 1.**
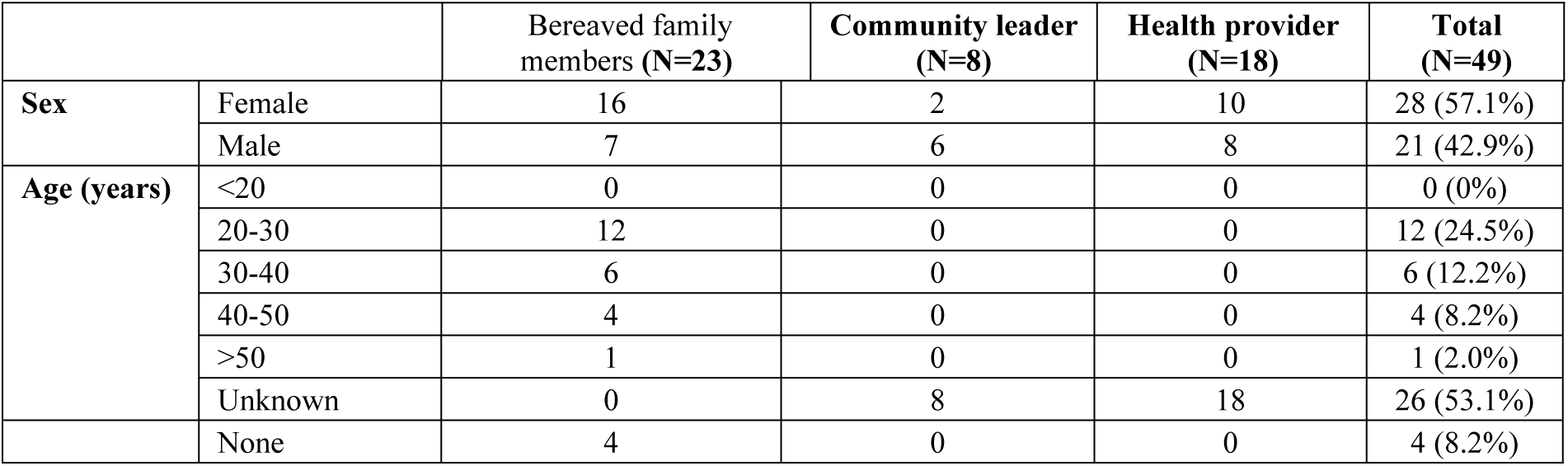
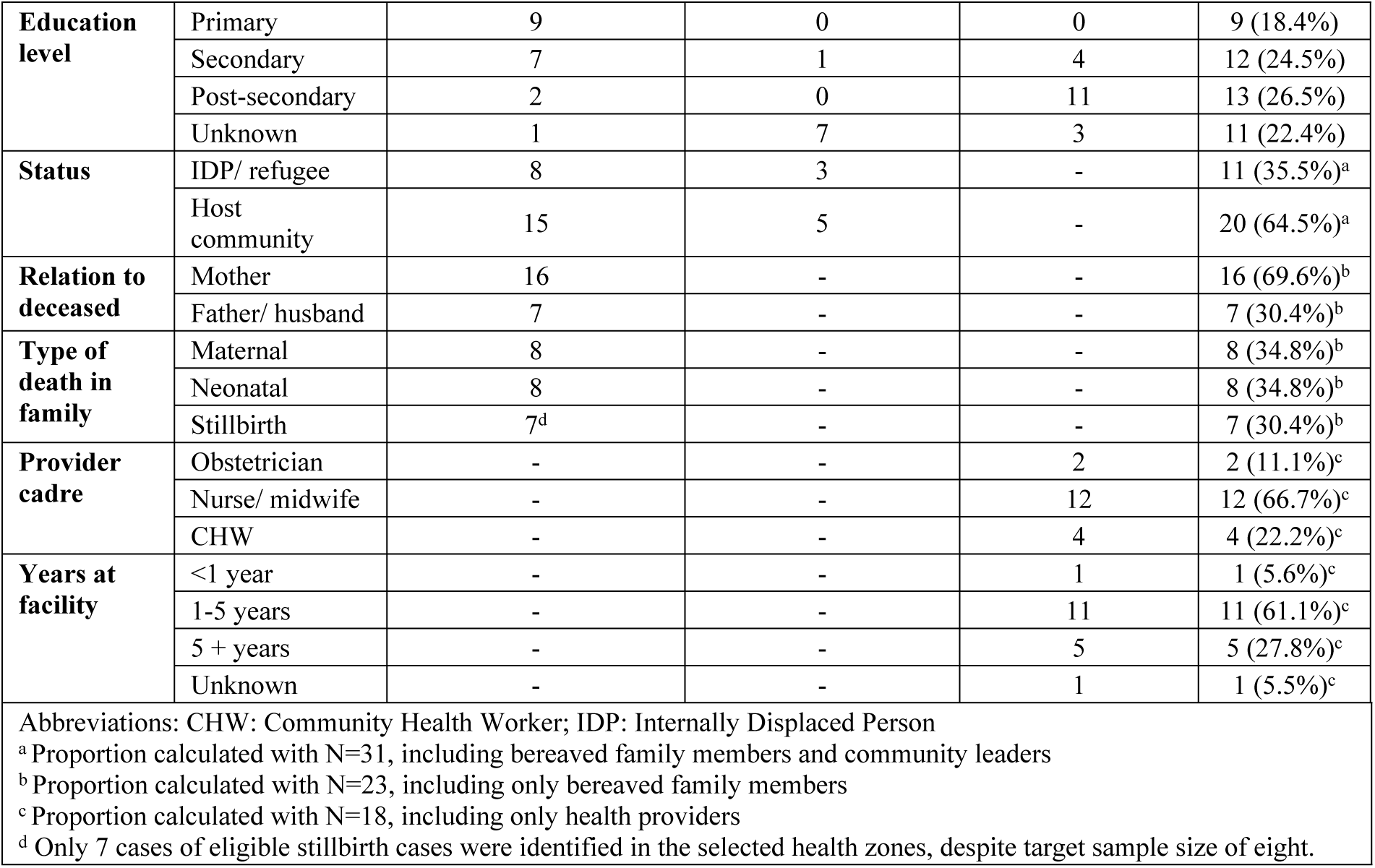
Participant characteristics.

### Socio-cultural practices surrounding maternal and perinatal deaths

Four key themes related to socio-cultural practices surrounding maternal and perinatal mortality were derived from the discussions with bereaved family members, community leaders, and health providers: i.) Burial practices; ii.) Mourning or bereavement traditions; iii.) Importance or rationale for bereavement practices or the lack thereof; iv.) Impact of insecurity on customs surrounding death.

#### i. Burial practices for maternal and perinatal deaths

Burial practices differed by type of death: maternal, neonatal, or stillbirth (Supplementary Materials Table 1). For maternal deaths, health providers shared that women were buried according to adult burial practices, including being placed in a coffin and buried. However, unique circumstances of deaths of pregnant women were discussed. The majority of participants shared how socio-cultural norms dictated that the woman must be buried separately from the fetus. For facility-based deaths, this required a cesarean section performed by a health provider to separate the fetus from the mother. However, for community-based maternal deaths, the chief or community leaders are tasked with the removal of the fetus from the woman’s womb. When separation of the fetus was not feasible within the community, health providers shared that communities buried deceased women in a grave next to a banana tree or burned banana leaves on the mother’s stomach to symbolize the separation of their two spirits.

> *“It was the women [of the community] who take her [deceased pregnant woman]. They’d go and dig a grave, but first they’d put a banana leaf on it. In case the autopsy wasn’t done, they’d put a banana leaf on the stomach, open it up and then bury it. Then they make another little grave, and this time they put a piece of a banana tree in it.”* – Community leader in South Kivu (8-D-3b-1)

A health provider also described other spiritual requirements:

> *“A woman who dies pregnant is treated in the same way as a woman who dies without giving birth. But custom dictates that we must bury her, and we cross the river before doing so. We do not bury her where we bury others; we must cross the river at all costs in order to bury her. That is what custom recommends… Her offspring’s spirit is left at the riverside…It’s the river that takes the offspring’s spirit, and so after crossing, she [the mother] can be buried.”* – Health provider in South Kivu (ID 8-D-4-1)

The burial of newborns and stillbirths were handled differently. In these cases, participants shared that men are not allowed to bury the corpse; burial must be handled by the mother or elderly women in the community. If the newborn death or stillbirth occurred at a health facility, participants shared that families are not authorized to take the corpse back to their community for burial; instead, it is buried in the vicinity of the health facility. If a newborn died within the community, it is put in a coffin and buried, typically in separate cemeteries since it did not “share life” with people in the community. For stillbirths, participants insisted that the burial must occur immediately after the event. In addition, many health providers added that local customs require that the stillbirth corpse be wrapped in his/her grandmother’s clothing and put in a box for burial. A health provider in South Kivu (ID 9-D-4-1) explained that “it’s often said ‘let’s not bury it with the mother’s clothes’ because it might take away the mother’s fertility.”

#### ii. Mourning or bereavement traditions

Participants also shared differences in mourning or bereavement practices (Table 2). For maternal deaths, bereaved family members, community leaders, and health providers shared that the bereavement period endures 2-4 days. Some bereaved family members specified that the first two days are devoted to mourning while the third and fourth days are focused on discussing the surviving children of the deceased mother. The payment of the deceased’s dowry is part of this discussion; participants shared that the husband must finish paying his wife’s dowry to support the needs of the children. In addition, a community leader and health provider explained that if the husband is suspected to have been the cause of the death (e.g., neglect, refusal to go to health facility, adultery), he may be obliged to pay his in-laws a fine of a goat.

**Table 2.** Sense-making of maternal and perinatal deaths.

| Themes | Maternal Deaths | Neonatal Deaths | Stillbirths |
| --- | --- | --- | --- |
| Witchcraft/Sorcery | Bereaved family members, community leaders, and health providers: deaths are linked to witchcraft. Examples:<br>Spells cast by the enemies, people the mother has been quarreling with, and in-laws will cause the death<br>Woman committed adultery<br>Blood transfusion using the blood of “unfaithful husbands” | Bereaved family members: Women who have experienced multiple neonatal deaths are accused of being a witch or of bewitchments | Health providers: Communities link stillbirths to witchcraft<br>Community leaders: Some say the woman's pregnancy was stolen by witchcraft |
| Religious beliefs | Bereaved family members: it comes from God | Community leaders: when a child dies, nobody killed him, it's God's will. | N/R |
| Blaming the mother | Bereaved family members: Even in her death, the mother may be blamed as being a recipient of witchcraft due to her adultery | Community leaders and health providers: mothers are often blamed and held responsible for negligence and non-adherence to counseling, health advice, or referrals shared by health providers<br>Community leaders: mothers will be looked down upon by their in-laws, especially if they have experienced several neonatal deaths and the community may justify separating the couple by saying she is unfit to run a household | Health providers: mothers are often blamed and held responsible for negligence and non-adherence to counseling, health advice, or referrals shared by health providers<br>Health providers: stillbirths are due to women’s consumption of indigenous products or adherence to societal taboos around cesarean deliveries |
| Blaming the husband | Bereaved family members: husbands are blamed for preventing their pregnant wives from going to the health facility or for neglecting their care<br>Bereaved family members: husbands may be accused of adultery | Community leaders: husbands may be accused of adultery while their wives were pregnant, causing complications for the baby. | Health providers: husbands’ attitude may be the cause of their wives' stillbirths |
| Blaming health providers | Bereaved family members, community leaders, and health providers: the community often accuse and blame the health providers.<br>When blamed, providers flee the health district out of fear of retribution from families<br>Intertribal conflict exacerbates blame when the health providers do not share the same tribe with the family of the deceased |  |  |
| Abbreviations: N/R: Not reported |  |  |  |

Bereaved family members explained that the parents of the deceased mother organize and pay for the funeral while the husband and children mourn. Expressions of solidarity and condolence gifts were common. Participants reported how family and community members would stay overnight to console the husband and children, often for extended periods of time to support the reception of others who would come to visit the family. If the family were religious, members of the church would accompany the corpse, pray, sing, hold mass, and collect offerings in the family’s honor. People within the community also expressed solidarity through visits to the family to mourn and console with the husband, and goats were commonly offered as a condolence gift to the family. A health provider shared that in some cultures a feast is also organized after 40 days of mourning.

Participants shared mixed practices regarding the mourning of newborns and stillbirths. Many participants explained that there is no bereavement period, ceremony, or mourning ritual for the loss of newborns and stillbirths. One community leader elaborated that in his culture, there is no mourning for the loss of the first child, only that of subsequent children. While participants recognized that there was no formal bereavement period for stillborn, some bereaved family members shared that people would sing en route to bury the stillbirth and sometimes family and other community members would visit the parents and console them.

For neonatal deaths, some participants shared examples of a short one-day bereavement period and others explained that the bereavement period depended on the family’s financial means. When traditions were practiced for a neonatal death, participants reported that the parents typically organized the bereavement period and received visits from their extended family and religious congregation. Other bereavement practices included consolation baths and the offering of goats in cases of families experiencing repeated loss (two or more). The death of one twin required the observance of specific bereavement traditions to protect the living sibling. Participants revealed that socio-cultural traditions forbid the surviving twin from seeing their deceased sibling, for doing so was believed to cause their death. For protection, tradition obliged the head of the family to perform the ritual of anointing the surviving twin with the blood of a slaughtered animal. The parents were also forced to hide their grief and mourn in private to shield the surviving sibling from the loss.

> *“If one of the twins were to die, according to our traditions here, you, the mother of the child, must not cry or even show that you have lost. Just be fine and act normal if you have one left. Because if you cry or mourn, the other child will die.”* – Health provider in South Kivu (ID 7-D-4-4)
>
> *“Either we slaughter a chicken or another animal. We anoint the survivor with the blood of this animal… The head of the family smears the blood on his hand as if he were blessing the child.”* – Health provider in South Kivu (7-D-4-1)

#### iii. Importance or rationale for bereavement practices

The importance or rationale for the specified bereavement practices for maternal and perinatal deaths were centered on culturally placed value of life and the pain endured by families after the loss of a mother or child. Participants felt strongly that an adult’s life should be commemorated and emphasized the necessity of recognizing and valuing a mother’s contribution to her family and community and coming together to support the family to cope with their pain.

> *“It’s important [to mourn] because she’s already a grown-up, everyone will cry, we’ll remember how we used to talk with her, they’ll eat, they’ll look at her photo, it’s important for her memory.”*-Mother who experienced perinatal death in South Kivu (9_D_3A_5)
>
> *“It is better to mourn for a pregnant woman because we have lived with her, we have shared her life, and if she dies while pregnant, we will mourn her deeply. And if a woman gives birth but her child dies, we will sympathize with her because she will have suffered for nothing, and the child belongs to everyone.”*-Mother who experienced perinatal death in South Kivu (9_D_3A_4)

Nonetheless, some participants acknowledged that while it is important to grieve for all people, grief for a child is not accorded the same socio-cultural significance as adults. Many participants shared how their society places little value on neonatal deaths and stillbirths, some recounting conditions of perceived societal value such as having teeth or contributing to a community.

> *“Everyone has their own way of doing things. In the case of stillbirth, there are those who do not consider it a loss; they bury the baby without mourning. But I think we should mourn a baby in the same way we mourn an adult. They are all human beings and require the same consideration.”* – Health provider in North Kivu (9-C-4-1)
>
> *“Let’s take the case of the mother who died leaving the child behind. Here, people gather immediately. All the neighboring villages, even those 10 km away, will come here. To attend the funeral, because it brings real sorrow, too much sorrow. It brings a lot of people together when a pregnant woman dies. Neonatal death is not as significant. People will say, “A child has died? Yes, it was the pregnancy of so-and-so’s wife.” Few people will come. But for this pregnant woman, even from 10 km away, people will come.”* – Health provider in South Kivu (9-D-4-2)
>
> *“If he didn’t have teeth yet, we don’t mourn him. They just go and bury him, and after the burial, everyone takes a bath to wash their hands, and then we tell everyone that there is no mourning, except that they can only come to comfort the mother.”* – Health provider in South Kivu (7-D-4-3)
>
> *“Well, the one you can’t grieve for is…stillborn. When people see that, they don’t have much to worry about. Unlike when a child is born… It brings a lot of sadness. You’ve already seen your child, and then after a short while, it dies; it’s very difficult to accept.”* – Health provider in North Kivu (9-C-4-3)

Some health providers also commented on families’ pain and loss in the aftermath of a neonatal death or stillbirth. They rationalized mourning and bereavement practices for newborns because “the pain of losing a newborn when the parents have seen it, is much greater than if it were a stillbirth.”

#### iv. Impact of insecurity on customs surrounding death

Many participants raised concerns about the ongoing crisis in Eastern DRC, highlighting its impact on customs surrounding death. Almost half of the bereaved family members of maternal and perinatal deaths (48%) expressed difficulties grieving and adhering to socio-cultural practices surrounding death within an insecure environment. Some shared that during escalations in violence and insecurity, corpses may be discarded without proper burial due to inability to obtain a coffin. Participating in burial practices, mourning, and/or bereavement traditions were also challenging, as population movement was often limited and safety in attending funerals and mourning with families was a concern.

> *“We used to do [celebrate at] funerals by playing music with speakers. But today you can’t do that if it’s 7pm and you’re still outside. That’s why it’s already changed. This war really, there are bandits, but they’ve become more numerous. You can’t stay out at night. We used to spend the whole night outside, lighting fires [to honor the death], but now it’s impossible.”* – Husband who experienced a maternal death in North Kivu (8-C-3a-2)
>
> *“You can’t mourn in the middle of a crackling bullet.”* – Mother who experienced a stillbirth in South Kivu (9-D-3a-5)
>
> *“Now when someone dies, we bury them and you flee. You, who are unsafe, how are you going to bury, how are you going to come together [to mourn]?”* - Community leader in South Kivu (8-D-3b-1)

### Sense-making of maternal and perinatal deaths

Identified socio-cultural practices surrounding maternal and perinatal deaths were deeply embedded in how communities constructed meaning around these events (Table 2). Witchcraft or sorcery was commonly cited as the cause of maternal and perinatal mortality. Participants described how maternal deaths were the result of spells cast by enemies of the mother or due to adulterous husbands. Neonatal deaths were also due to bewitchment or the mothers themselves being a witch and stillbirths were conceptualized as a woman’s pregnancy “stolen by witchcraft.”

> *“If she was at home, people say she was either bewitched or she was not properly cared for. People say things like that; there’s no shortage of comments like that. But if she died while in the hospital, there’s really no problem.”-Community health worker in North Kivu (9_C_4_3_)*
>
> *“ Here, if it becomes a habit to lose children every time, they might even call you a witch, which is why you lose children often.” - Mother who experienced stillbirth in North Kivu (8_c_3a_5)*

The interpretation of deaths through a religious lens was referenced far less frequently. However, a couple of bereaved family members and leaders shared that maternal and neonatal deaths were “God’s will.”

Blame was a common thread woven in societal norms to rationalize maternal and perinatal deaths. Blaming the woman, even on her deathbed, was recurrently articulated by participants. For maternal deaths, women were blamed for adultery. For neonatal deaths and stillbirths, women were blamed for negligence and non-adherence to counseling, health advice, or referrals requested by health providers. Several community leaders further noted that mothers were often stigmatized following neonatal deaths, especially by their in-laws, and that in some cases the community may justify marital separation on the grounds that the woman is “unfit.” Health providers also blamed women, claiming that adherence to cultural norms and traditional practices (i.e., consumption of indigenous products, taboo around cesarean delivery) caused their stillbirth.

> *“This traditional product that these women take is eggs. In our jargon, they call it “Katulumula,” which is the name of these eggs. They break them and drink them in order to conceive quickly. So, they take it without knowing the dosage and without knowing what effect it has on the child… And then they come to the hospital, but it has a harmful effect on them.”* – Health provider in South Kivu (ID 8-D-4-1)
>
> *“There was a witch doctor in this village who said that mothers had to undergo surgery because of the poor shape of their pelvises and that he was able to fix it. He had medicine for it. So now, they [women in the community] started going to see this man who was tricking them. He would treat two women for the price of a goat.”* – Health provider in South Kivu (ID 7-D-4-2)
>
> *“We, the health providers, give information, but there are others who counteract this information. ‘God said, God revealed, he showed this…’ In our customs here, a woman who does not give birth vaginally is not considered a woman. If you always have to open the abdomen to remove the child, you are not a worthy woman. A woman knows how to push the child out, and the child must pass through the vagina, but since she says to herself, ‘I am always discredited by my family, I must give birth vaginally,’ she will remain in labor at home. When it becomes complicated and she is exhausted, that is when you will see her being carried on a stretcher to the facility.”* – Health provider in North Kivu (ID 8-C-4-3)

Communities also made sense of maternal and perinatal deaths through the blame of husbands and health providers. Bereaved family members explained that husbands may be blamed for adultery, neglect, or preventing their wives from attaining health care. Participants also reported that communities frequently accused and blamed heath providers for the deaths of mothers and babies, at times leading health providers to flee the health district out of fear of retaliation from bereaved families. Additionally, health providers revealed that intertribal tensions in the region contributed to heightened blame toward health providers, particularly when they do not belong to the same culture or ethnic group as the deceased’s family.

> *“If someone already knows that this doctor hid the truth [cause of neonatal death] from me and I only found out later, trust declines, and you’ll see later, ah, they hid the truth from me, even if I have a sick person, I won’t go there anymore… I’m not going to get treatment here anymore”* – Mother who experienced stillbirth in North Kivu (8_c_3a_5)
>
> *“We have experienced many periods of war here… and even more inter-ethnic wars… Since it was a certain ethnic group that delivered the baby and both the child and the mother died, we began to attribute this to ethnic groups, to tribes…”* – Health provider in North Kivu (ID 8-C-4-3)

### Implications for Maternal and Perinatal Death Surveillance and Response

Socio-cultural practices and community sense-making of maternal and perinatal deaths had significant implications on the functionality of an MPDSR system. Limited disclosure within communities impedes case identification and mortality reporting to the health sector. Participants reported that maternal deaths were more often disclosed and reported than perinatal deaths. However, community-based maternal deaths were not spoken of or reported, in avoidance of blame or punishment. Moreover, the attribution of unexplained maternal and perinatal deaths to witchcraft limited disclosure and reporting and some socio-cultural practices and customs even forbid it. In the aftermath of a death, many participants shared that family members do not have the strength to disclose the significant loss of a mother or child and/or may want to forget the event. Additionally, perinatal deaths were often not disclosed or reported due to their low valuation in society. Overall, participants described limited awareness of the importance of reporting deaths that occur within the community. One mother who lost a newborn (ID 8-D-3A-2) shared, *“So, a death that you don’t know how to explain, you’ll keep it inside. Those who come when you get home to the neighborhood will talk about how the death happened, but when you yourself don’t know what happened [you keep it inside]…”*

> *“ Yes, there are deaths that we cannot announce, and that is when we do not know if evidence has arrived, how we are going to write that someone has died when you do not have evidence of witchcraft for example”—health provider in South Kivu (7-D-3B-1)*.

## Discussion

Within MPDSR, scholarly and programmatic publications have increasingly included attention to community engagement strategies and social pressures influencing health worker behaviors, particularly the blame dynamics that undermine death review processes (16,20–22,29,30). However, less attention has focused on how socio-cultural practices shape whether and how deaths are disclosed and reported. This study addresses that gap by examining how culturally embedded practices surrounding death influence case identification and reporting in two conflict-affected health zones in Eastern DRC.

Drawing on social constructionist perspectives (31,32), our findings illustrate how stillbirth, neonatal death, and maternal death are understood through distinct cultural frameworks that shape both emotional responses and reporting practices. Stillbirths and neonatal deaths were frequently viewed as spiritually or socially less significant than maternal death because the child had not yet contributed to the community or shared life with others. This cultural valuation underpins shortened or absent mourning rituals, immediate burial, and limited acknowledgment by extended family or community members. In contrast, maternal deaths were seen as highly significant events requiring multi-day bereavement, community mobilization, and symbolic practices to separate the spirits of mother and fetus. Cultural constructions of death are intertwined with local beliefs about causality, including witchcraft and spiritual destiny, that shape both mourning practices and reporting behavior.

Critical medical anthropology emphasizes the importance of understanding how institutional structures and political dynamics can intersect with cultural norms to influence reporting (33). Participants reported low awareness of MPDSR processes, lack of clarity on responsibilities for community-level reporting, and persistent taboos that limit discussion of maternal and perinatal deaths. Participants described a context in which insecurity and conflict dynamics disrupt customary burial and mourning practices, restrict movement, and intensify fear, echoing findings from other contexts where interpretations of misfortune are co-produced by communities and influence health system engagement (34–38).

To improve community-based identification and reporting of maternal and perinatal deaths, MPDSR guidelines and procedures (12) should be adapted to consider local values, norms, and socio-cultural practices that may influence implementation. Figure 1 outlines key considerations to culturally optimize MPDSR in Eastern DRC.

**Figure 1.**
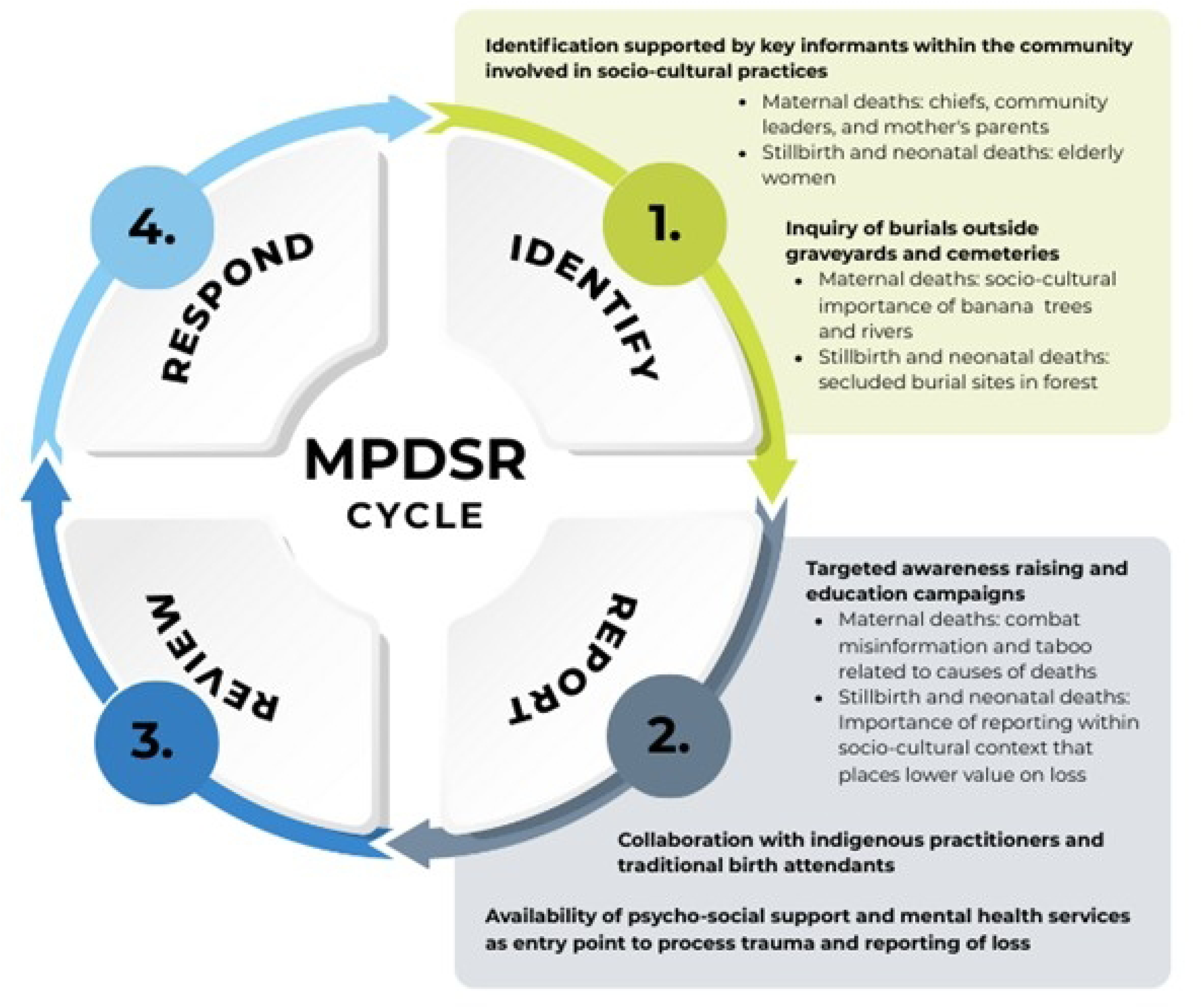
Culturally optimized MPDSR system to improve identification and reporting of community-based maternal and perinatal deaths

First, WHO guidance on MPDSR describes the use of community health worker cadre to identify community-based maternal and perinatal deaths (12). Our findings suggest that case identification should be supported by key informants within the community that engage in socio-cultural practices surrounding death. For maternal deaths in Eastern DRC, village chiefs, community leaders, and the parents of the deceased mother are key focal points given their integral involvement in burial and bereavement traditions. Due to the limited disclosure around neonatal deaths and stillbirths, elderly women within communities who are most often tasked with the burial, should be considered as an important actor with intimate knowledge of these losses.

Second, identification of these cases within the community should also consider inquiries about their burial location, which due to the cultural sensitivity of losing mothers and children, are typically not with the general population. The socio-cultural significance of burial sites near banana trees and rivers is noteworthy for maternal deaths. For perinatal deaths, cases may be identified within separate secluded burial sites often within the forest.

Finally, efforts to identify maternal and perinatal death cases with culturally appropriate key informants and burial sites should be complemented with targeted efforts to improve disclosure and death reporting. Awareness raising and education campaigns within communities are needed to improve understanding around the importance of MPDSR and directly address prevalent socio-cultural beliefs that impact death reporting to the health sector. For example, education campaigns that address misinformation around the cause of maternal deaths in Eastern DRC may combat the taboo and fear around reporting deaths. Collaboration with traditional healers, traditional birth attendants, and religious leaders may also improve reporting.

Past research and program experiences support similar culturally responsive models for public health surveillance. For example, during the Ebola outbreaks in West Africa and DRC, social scientists played a critical role in designing interventions that respected traditional burial practices while ensuring safe procedures to prevent disease transmission; however, where political instability and institutional neglect had eroded community trust, even culturally adapted responses struggled with concealment and resistance (39–42). Similarly, culturally adapted mental health interventions have drawn on community norms, language, and traditional forms of support. In the DRC and neighboring countries, mental health programs have worked with lay counselors and local elders to address psychological distress using familiar, acceptable approaches such as faith-based counseling, storytelling, or group rituals (43–45). What these efforts share is a recognition that participatory approaches which shift power toward communities are more likely to produce sustainable health system changes (46). Similar dynamics have been documented in maternal health programs (47), underscoring the importance of engaging communities in contextual adaptation of MPDSR systems.

## Strengths and limitations

As a phenomenological qualitative study, findings represent the experiences and perceptions of a specific population in two purposively selected health zones in Eastern DRC and are not designed to be generalizable. This study benefits from a methodology that captured diverse perspectives and triangulated insights from bereaved family members of maternal and perinatal deaths, as well as community leaders and health providers. Conducting interviews in participants’ preferred language (French or Swahili) and in locations chosen by participants themselves likely contributed to data quality. Even so, the sensitivity of the subject matter may have introduced social desirability bias, and recruitment through CHW networks and facilities that had reported deaths may have skewed participation toward families and providers with greater health system engagement. Additionally, transcription and translation of interviews may have led to some loss of nuance. Despite these limitations, the study provides valuable insights into how burial practices, mourning customs, and local sense-making processes influence the reporting of maternal and perinatal deaths. By identifying specific community actors, ritual practices, and culturally meaningful burial sites in participating health zones, findings offer actionable entry points for strengthening MPDSR in Eastern DRC.

## Conclusion

Effective MPDSR requires moving beyond implementation of standardized reporting and review frameworks, to understand the socio-cultural dynamics that shape whether deaths are discussed and how communities interact with formal health structures. Our findings show how cultural beliefs, social norms, mourning practices, and community trust influence whether maternal and perinatal deaths are identified and reported. National MPDSR guidelines and policies must therefore be adapted to reflect local cultural norms, incorporating community-based actors into case identification processes and demonstrating genuine respect for local mourning practices and social dynamics that may inhibit disclosure. Investing in culturally responsive MPDSR is not an optional consideration but an essential condition for generating reliable mortality data. As the global health community works to reduce preventable maternal and newborn deaths, this study underscores the limitations of current maternal and perinatal mortality data sources and importance of integrating social and cultural analysis into surveillance system design and health program planning,

## Data Availability

Due to the sensitivity of the study, qualitative data is available from the corresponding author on reasonable request and signature of a data sharing agreement.

## Declarations

### Ethics approval and consent to participate

The study protocol was reviewed and approved by the Johns Hopkins Bloomberg School of Public Health institutional review board (IRB No.: 21935) and the Universite Catholique de Bukavu Ecole de Sante Publique ethical review committee (UCB/CIES/NC/022/2022). All study procedures have conformed to the principles embodied in the Declaration of Helsinki. Written informed consent was obtained in-person from all respondents in advance of data collection.

### Consent for publication

Not applicable.

### Availability of data and materials

Qualitative data is available from the corresponding author on reasonable request and signature of a data sharing agreement.

### Author Contributions

MM, CCM, HT, EG, PM, and GBB designed the study and data collection tools.

MM, CCM, EG, GN, and CCZ led the data collection with the support of GBB and PM.

MM and CCM analyzed the data.

MM, CCM, HT, SE, PM, and GBB prepared the initial manuscript draft.

All co-authors reviewed and contributed to the final version of the manuscript.

## Acknowledgements

We thank all the participants who shared their stories and experiences with maternal and perinatal deaths in their communities. We thank the research team from the Regional School of Health at the Universite Catholique de Bukavu, as well as colleagues from the Provincial Health Division of North and South Kivu, Directorate of Studies and Planning of the Ministry of Health, National Program for Reproductive Health of the Ministry of Health of the DRC, and the International Rescue Committee DRC office who supported our research.

